# Lessons learned from operationalizing the integration of nutrition-specific and nutrition-sensitive interventions in rural Ethiopia

**DOI:** 10.1101/2023.08.11.23293982

**Authors:** Girmay Ayana Mersha, Eshetu Zerihun Tariku, Wanzahun Godana Boynito, Meseret Woldeyohaness, Birhanu Wodajo, Tadese Kebebe, Stefaan De Henauw, Souheila Abbeddou

**Affiliations:** Food Science and Nutrition Research Directorate, Ethiopian Public Health Institute, Ethiopia; Public Health Nutrition, Department of Public Health and Primary Care, Ghent University, Belgium; School of Public Health, Arba Minch University, Arba Minch, Ethiopia

**Keywords:** Community health, agriculture, multisectoral, nutrition, Ethiopia

## Abstract

**Background:** Undernutrition reduction requires coordinated efforts across sectors to address its causes. A multisectoral approach is important in diagnosing the problem and identifying solutions that would be implemented across different sectors.

The study aims to explore the experience of health and agriculture extension workers in integrating nutrition services provided to households with children under two years of age at community level.

**Methods:** A qualitative study has been conducted in agrarian areas of Ethiopia in 2021 at the end of a large intervention program to reduce stunting. In total, 28 key informant interviews were conducted with health- and agriculture-extension workers and mothers. A framework analysis approach was applied to manage and analyze data using NVivo version 12 software.

**Results:** Community-level joint intervention is a feasible approach to integrating nutrition services. Farm gardening and cooking demonstrations were practiced jointly with extension workers and households. Because of service integration, extension workers perceived an improved father’s role in supporting mothers in childcaring and feeding nutritious diets to children, and decreased severe cases of undernutrition. Integration of health and agriculture sectors for nutrition intervention was challenged by the high workload on extension workers, poor supervision and leadership commitment, lack of appropriate agricultural inputs, and absence of clarity on sector-specific roles. In some areas nutrition services are not owned by the health and agriculture sectors, and it was overlooked.

**Conclusion:** Integrating existing community-level platforms are key entry points to address undernutrition and promote key agriculture and health interventions in Ethiopia. Empowering extension workers through training, monitoring, and supervision is part of ensuring the sustainability of integration between sectors.

## Introduction

Undernutrition including stunting, wasting, underweight and micronutrient deficiencies plagued populations in low- and middle-income countries for decades and remains one of the most pressing health problems. Young children and women of reproductive age are particularly vulnerable because of their increased nutrient requirements, and, in case of women, their lower social status, which limits their access to nutrient-rich foods [1]. The causes of undernutrition are complex and affected by factors acting at different causal hierarchical levels [2]. The political, economy, health, education, agriculture and food systems, water and sanitation, and the environment are all determinants of maternal and child nutrition and health [3]. Undernutrition reduction requires coordinated efforts across sectors to address its underlying causes [4, 5]. Addressing the problem of undernutrition needs the collaboration of multiple sectors and a variety of stakeholders in governments, non-government organizations, and the private sector [6]. Undernutrition reduction requires a departure from a health system-centered approach of provision of nutritional services and nutrition-specific interventions only, to an integrated approach that encompasses nutrition-sensitive, food systems-based approaches[7].

The framework of actions proposed by the UNICEF and the Lancet nutrition series recognizes the needs for multiple actions that are at different levels from immediate, underlying to distal [2, 8]. Interventions targeting the improvement of maternal and child nutrition and health act on interlinked factors which influence each other. Nutrition-specific interventions address the immediate causes of undernutrition and nutrition-sensitive interventions focus on the underlying causes of good nutrition [9]. Both nutrition-sensitive and specific interventions require an enabling environment, that encompasses actions addressing basic determinants of undernutrition. Multiple actions to improve nutrition were undertaken over the recent years, the most prominent is Scaling Up Nutrition which clearly underscores the necessity of actions among all the stakeholders [10]. This approach puts nutrition at the core of each sector plan and institutional effort.

However, systematic coordination between different sectors to achieve the objectives of improved maternal and child nutrition is problematic, given the bureaucratic barriers that characterize the administrative division of responsibilities among sectors. All sectors are expected to be clear about what they can contribute to resolving malnutrition in a particular context specific to their sector role. Agriculture, as an example of nutrition-sensitive intervention, can potentially increase food availability, which would directly improve the dietary accessibly leading to improved nutritional status of targeted populations. However, the multiple factors that influence nutrition, the social dynamics within households that can result in unexpected allocations of resources, and the many indirect ways that agriculture may impact nutrition, require purposeful planning and careful consideration of how exactly program inputs will lead to nutritional improvements. This complex relationships has consequently lead to limited impact of nutrition intervention, because other determinants are not put in place at different levels.

Ethiopia is one of the countries with the highest rate of undernutrition in sub-Saharan Africa. The proportions of stunting, underweight and wasting among children under age of five years are respectively 38%, 24% and 10% in Ethiopia [11]. Multiple efforts have been made to implement the government commitment towards ending undernutrition. To reach this goal, Ethiopia has demonstrated a strong policy commitment to nutrition through the development of a multisectoral food and nutrition policy, which highlights the need to accelerate multi-sectoral approaches [12]. Political willingness is an important factor that helps to implement effective nutrition interventions [10]. However, the Ethiopian situation remains in its debuts with poorly synchronized sectors of food, nutrition, agriculture, and health to prevent the problem of under- nutrition. In Ethiopia, the presence of health and agriculture extension systems and frontline workers at the community level is a good framework that facilitates the synergetic linkage of nutrition within health and agriculture services [13] Additionally, existing multisectoral nutrition coordination structure at all levels, and the willingness of international and national development partners to support multisectoral nutrition interventions are favorable conditions on the ground [14]. Ethiopia has implemented multiple nutrition-specific and nutrition-sensitive programs such as the Ethiopian national nutrition program, and the Seqota declaration program, that have required the development of structures and processes that facilitate multisectoral coordination [15]. But, collaborations between the health and agriculture sectors are suboptimal at community levels [16]. The multi-sectoral nutrition programs and interventions lack strong coordination that brings on board all the relevant stakeholders.

Based on these facts, Ethiopia designed different intervention modalities whose objective is to strengthen community-level multisectoral nutrition interventions. In this context, the Sustainable Undernutrition Reduction program in Ethiopia (SURE) was developed and implemented between 2016 and 2020 in rural regions of Ethiopia. The program utilizes the potentials of the Agricultural Development Agents (DAs) and the Health Extension Workers (HEWs) for the promotion of improved agricultural practices and utilization of high nutrient products to promote dietary diversification among rural mothers and children. At present, there is no study that has analyzed the experience and perception of the health (HEWs) and agriculture extension workers (AEWs) in integrating nutrition services provision at the community level in rural Ethiopia. Understanding frontline workers’ practices and experience is crucial for identifying appropriate measures to sustain nutrition improvements [9]. This qualitative study aims at appraising the understanding, the knowledge and perception of those women and men who are the most in contact with the communities in rural Ethiopia and who were responsible to implement the SURE program.

## Methods

### Study Setting

The Ethiopian Ministry of Health and the Ministry of Agriculture led the implementation of the SURE program whose aim was to reduce stunting by improving complementary feeding practices and dietary diversity of young children through a process of improving local agricultural productivity, food accessibility, food consumption diversity and nutritional status of young children and women [27]. SURE, was implemented between 2016 and 2020 in four agrarian regions (Oromia, Amhara, Southern Nations, Nationalities, and Peoples’ Region [SNNPR], and Tigray). The program package included promoting diversified agriculture, promoting infant and young child feeding (IYCF) practices, empowering women’s decision-making related to agriculture, diversified food intake, and health, and enhancing food security and water, sanitation, and hygiene practices (WASH). The SURE intervention districts were selected because of the high food insecurity and stunting prevalence in four regions out of the nine total regions in Ethiopia. Details of the SURE intervention program have been published elsewhere [17].

Briefly, a total of 36 districts (Woreda as locally called) were selected and assigned to the SURE intervention. Comparison control districts were selected in equal number (36 districts) in the same four agrarian regional strata as for intervention districts. Districts were roughly matched based on stunting prevalence tertiles (low, medium or high) and food insecurity scores. Eligible households are households with at least one child under the age of 2 years. The intervention started in 2016 and included 4299 households, 2146 households from 144 Kebeles (smallest administrative unit) were included in the intervention group and 2153 from 144 Kebeles in the control group. After baseline data collection, and all households received counselling at household and community levels. Additionally, 10 % of households with children aged under two years in the intervention regions (they can be enrolled or not in the larger SURE intervention) were 1) provided improved seed variety and chicken through AEWs and 2) trained through demonstration gardens at farmer training centers.

The SURE program was delivered through: 1) interpersonal contacts to provide counselling on IYCF and nutrition-sensitive agriculture advice to mothers and fathers of children under 24 months, inclusive of pregnant women and fathers-to-be, jointly delivered by the local HEWs and AEWs during routine household visits; 2) men’s and women’s group dialogues targeting all men and all women in a given community network, facilitated also by the local HEWs and AEWs, and 3) media campaign to reinforce IYCF and dietary diversity messages.

### Study design

A phenomenological qualitative study design was conducted among beneficiaries (mothers) and implementers (HEWs and AEWs) of the SURE program in the intervention communities. One Kebele per district from the intervention group was randomly selected to contribute in the qualitative study.

### Study period

This qualitative study was conducted after four years of intervention. The data were collected at the community level in selected districts of Amhara, Oromia, and SNNPR between January and February 2021. Data collection was discontinued in Tigray because of the war.

### Recruitment of participants

All SURE intervention districts and Kebeles in the three agrarian regions of Ethiopia were eligible for inclusion. The recruitment of participants was done purposely to obtain maximum variation of information. The HEWs and AEWs as service providers of the intervention packages, and mothers who were enrolled in the SURE intervention as direct beneficiaries were invited to participate in this study. The HEWs and AEWs were eligible to participate in this study if they received the SURE program training in 2016 and have provided the service as per their training in their respective Kebele for a minimum of two years. Mothers should be residents of the Kebele, would have had a child 0-24 months old at the time the intervention started, and received joint health and agriculture counseling as described in the study setting.

### Data collection and tools

Data were collected using key informant interviews (KIIs) at the community level from a total of 28 eligible respondents including HEWs, AEWs and mothers. Participants were asked about their understanding of nutrition, quality of service provided and quality of nutrition education specific to the SURE program, the challenges of implementing nutrition programs jointly, and how the program could be sustained for a better nutrition service provision (Supplementary material 1).

Participants took part in a face-to-face interview. A topic guide was developed by the research team and training on how to conduct KIIs was provided. The interview began with demographic questions about schooling, experience, and professional background. All the interviews were conducted in a private space and were audio recorded. Data was collected in the local languages (Amharic and Oromifa). Audio recordings were transcribed and translated into English. All transcribed data was anonymized.

### Data Analysis

A framework analysis approach was used to manage and analyze the data. Framework approach provides clear steps to follow and produces structured outputs of summarized data that can be used with a variety of data sources, including interviews, focus groups, and documents [18]. The steps include familiarization (becoming familiar with the content of the data), Identification of thematic areas (identifying a thematic framework, which is a set of predetermined themes or categories that are relevant to the research question), Indexing (systematically coding the data to identify content related to each theme or category), and Interpretation (identifying patterns and relationships across the data, and interpreting what these patterns and relationships mean in relation to the research question). Qualitative data analysis software NVIVO version 12 software was used in coding and constructing thematic areas from the collected data. The transcripts were coded into specific themes. The themes were identified and coded based on the topic guide as nutrition understanding, achievements, opportunities, and way forwards for a successful implementation of nutrition-specific and nutrition-sensitive programs and sustainability.

### Ethical considerations

Ethical approval was obtained from the research ethic committees both at the University Hospital of Ghent, Belgium (protocol number BC-08862), and the Ethiopian Public Health Institute, Ethiopia (protocol number EPHI-IRB-306-2020). The qualitative study included recipients from the same cohort (participating mothers from the intervention group), in addition to health extension workers from the intervention Kebeles. Written informed consents, which included permission to be recorded using a digital audio recorder, were obtained from participants before interviews. Further administrative permissions were granted by local government bureaus in all regions.

## Results

### Demographic characteristics of study participants

From the total of twenty-eight key informants who participated in the study, 43%, 39% and 18% were from Oromia, Amhara and SNNPR regions, respectively. The proportion of HEWs, AEWs and mothers were almost similar. Totally 58 % of the extension workers were femaleand 73% had at least five years of work experience. Sixty-seven percent of the study participants have completed college and above.

**Table 1:**
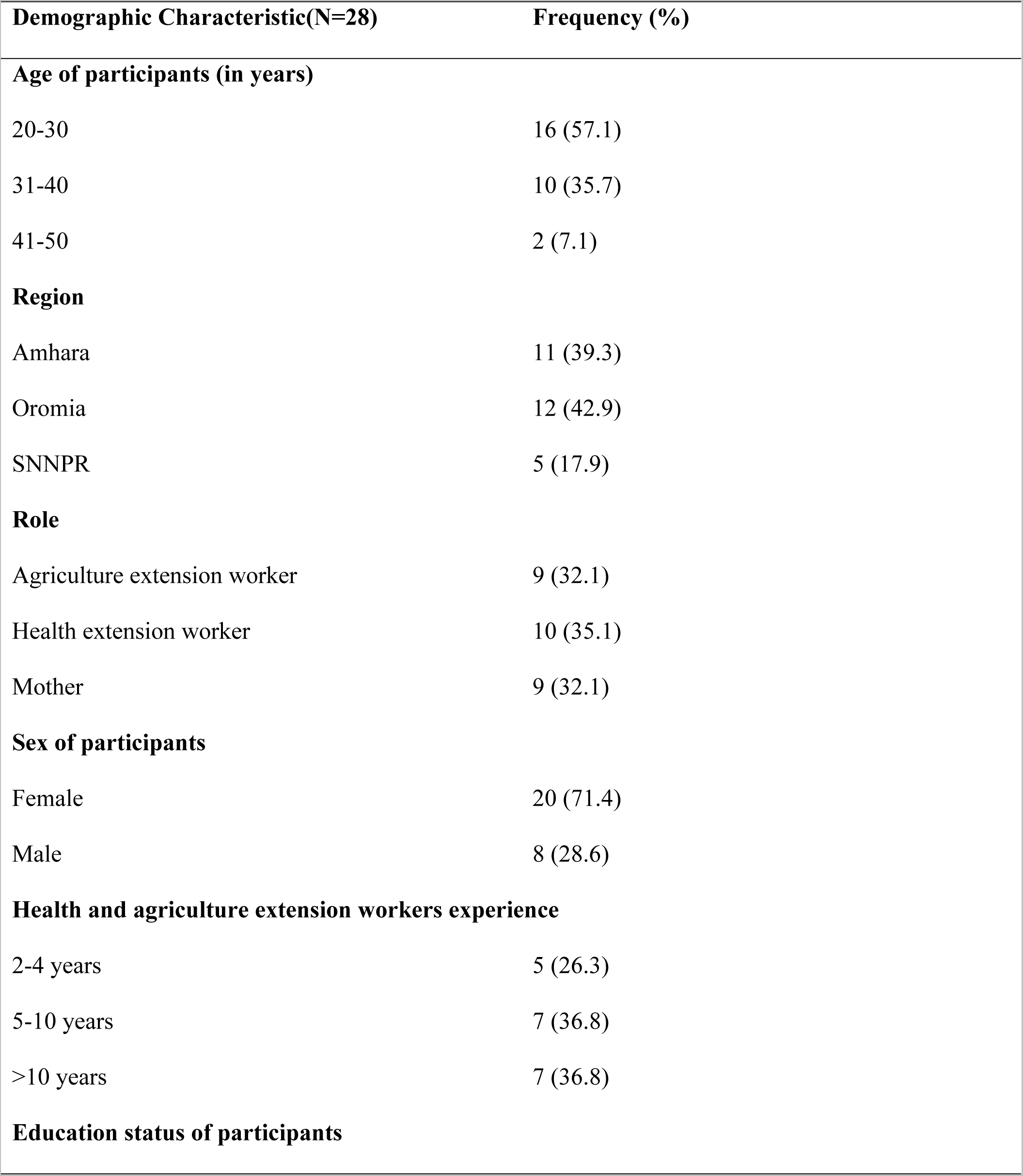
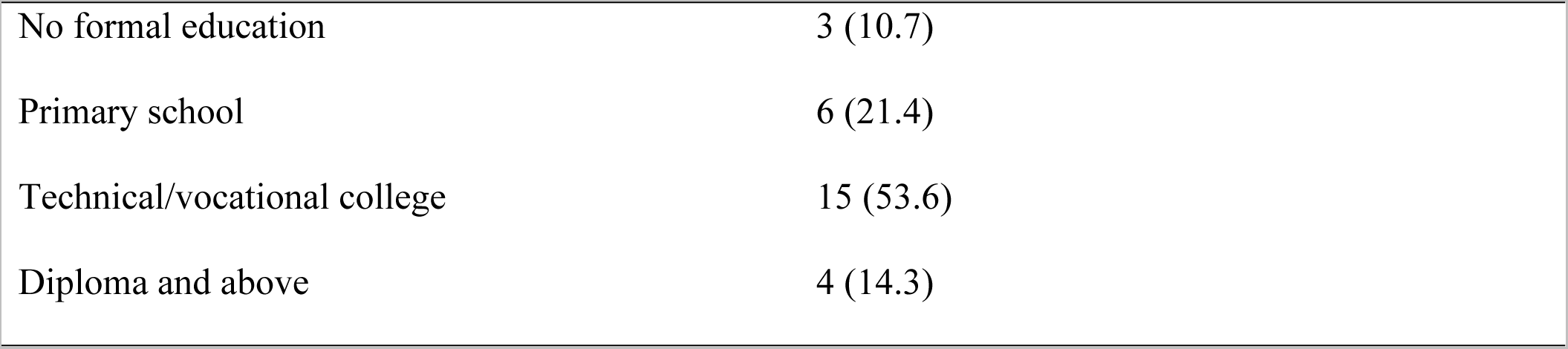
Demographic characteristics of the qualitative study participants, Ethiopia.

| Demographic Characteristic(N=28) | Frequency (%) |
| --- | --- |
| **Age of participants (in years)** |  |
| 20-30 | 16 (57.1) |
| 31-40 | 10 (35.7) |
| 41-50 | 2 (7.1) |
| **Region** |  |
| Amhara | 11 (39.3) |
| Oromia | 12 (42.9) |
| SNNPR | 5 (17.9) |
| **Role** |  |
| Agriculture extension worker | 9 (32.1) |
| Health extension worker | 10 (35.1) |
| Mother | 9 (32.1) |
| **Sex of participants** |  |
| Female | 20 (71.4) |
| Male | 8 (28.6) |
| **Health and agriculture extension workers experience** |  |
| 2-4 years | 5 (26.3) |
| 5-10 years | 7 (36.8) |
| >10 years | 7 (36.8) |
| **Education status of participants** |  |
| No formal education | 3 (10.7) |
| Primary school | 6 (21.4) |
| Technical/vocational college | 15 (53.6) |
| Diploma and above | 4 (14.3) |

### Major causes of undernutrition

The causes of undernutrition are seasonal and context-specific. The key informants perceived that, in their community, undernutrition was due to a shortage of land to produce adequate food to sell or to consume for a rapidly growing population. The severity of the problem increases when it is coupled with poor distribution of agricultural supplies, and pest infestation that affect productivity.

> *Absence of enough farmland is the cause of undernutrition. Since the community is large in number, farmland is not enough. on average the farmland owned by the farmers is about 0.375 up to 0.5 hectare. AEW,SNNPR.*

The problem of land and water scarcity were identified as the main cause of undernutrition in the community in Oromia region. The small plot of land is challenged by a shortage of water and agricultural inputs.

> *The lack of adequate farming* place *also another problem because it is difficult to produce enough food for family consumption using one farming place and leads to poverty. Moreover, lack of adequate rain, lack of irrigation service, and the fertilizer supply is not as required in our kebele, which may lead to poverty. HEW, Oromia*.

Access to fruits and vegetables is important to improve food security and undernutrition reduction. Frequent drought and absence of irrigation facilities to farm during the dry season are among the causes of undernutrition that undermine the efforts of AEWs and households to improve food and nutrition interventions.

> *Malnutrition is mainly due to insufficient production of fruits and vegetables due to drought in this area. When the husbands and wives agree to produce in their home garden, it will vanish, because there is no irrigation. Due to this reason our community failed to produce fruits and vegetables and give less emphasis. HEW, Oromia*.

War and civil unrest affect access to- and availability of foods through destructing market linkage. Instability compromises the support of HEWS and AEWs to the community. An extension worker indicated that the causes of undernutrition are related to lack of hygiene and sanitation, diseases and infections, inflation and war that displaced people from their residential areas and hindered them from producing food.

> *The cause of under* nutrition *are lack of hygiene, disease, war, and such things are the cause of under nutrition in addition to the above, there are food security issues. For instance, there is a lack of milk for children, they can’t access enough milk. There is also limited access to eggs. HEW, SNNPR*.

> *Now in some* places*, based on market inflation, some people become incapable of buying something, as a result, the child may develop malnutrition. For example, when children need milk, they may fail to give it. As a child fails to get balanced food when they need and passed that time, they may go into malnutrition. Some households are economically incapable to care for their child, there is a household that relies on the market business for leading the family, such a woman gave other foods to their child before six months and leave the child at home because they can’t stay home and breastfeed their child regularly. HEW, Oromia*.

### Challenges of integrating health and agriculture interventions in Ethiopia

Many challenges have been reported by extension workers in the implementation of intersectoral interventions. High workload, conflict, shortage of agricultural inputs, limited training and poor supportive supervision and COVID-19 pandemic were the most frequently reported challenges (Figure 1).

**Figure 1:**
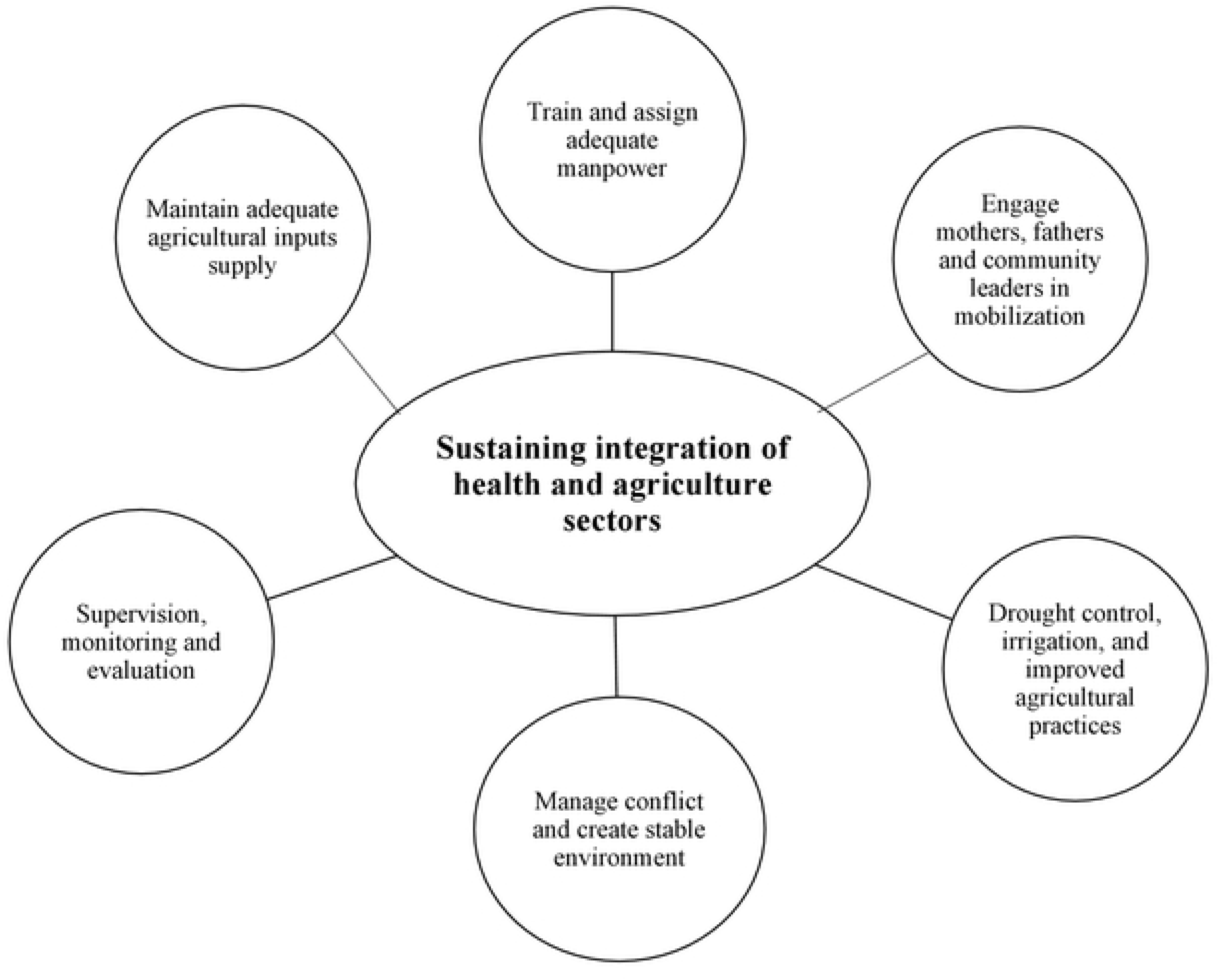
Sustaining the integration of agriculture and health sectors for reduction of undernutrition in Ethiopia

### Intervention in the midst of the war in Ethiopia

Ethiopia is facing civil unrest in the last four years and the project intervention has been affected. Conflict was a challenge of service delivery at community level, which resulted in destruction of facilities, damages of means, and loss of income by restricting people’s movement and targeted attacks on workers.

> *The current situation is* overly *sensitive. It is now three years since the conflict began. Every day there are riots, there is something. So, people think it is turning to something else. At the beginning of the SURE program, the expert was there to support them every day, and the people were getting education and had been given training. It was good to take care of that and practice. But now people are abandoning everything and entering politics. ’’Who will die today? Who will live today?’ They are living in fear. In this regard, political stability is a bit difficult. AEW,SNNPR*.

Conflict and the COVID pandemic have also compromised the supportive supervision and monitoring activities by restricting people’s movement from regional and zonal levels to front line workers.

> *… So, this team has* met *frequently, and minutes have been kept on our discussion. This meeting was previously held every week, then turns to every month. As I said earlier, recently we have less engagement due to varied factors and we do not have minutes. We fail to gather the participants to give different training as this contradicts the covid-19 restrictive measures. HEW, Oromia*.

### Shortage of agricultural inputs and material for workshop training

The availability of agricultural inputs for gardening and cooking demonstrations at community level was a challenge and participants were not interested in bringing their own food and material for demonstration. The project gives agricultural inputs to only 10% of the total population, but the demand for agriculture inputs was high and the community perceived that the project does not treat people equally.

> *We are very* much *interested in the work (for example a demonstration of food preparation). For this demonstration, if you asked the farmers to bring the ingredients/inputs (like egg, kale, cereals, ….) from their home they will lose their interest. It is difficult to bring by yourself too. The issue is the problem of access to input for the demonstration. AEW, Oromia*.

### Workload and ownership of the program

Nutrition activities are not a priority agenda in all sectors equally. The ownership of nutrition is not clearly defined, and extension experts assume an additional assignment is beyond their regular tasks. There is a lack of awareness and commitment by AEWs to implement nutrition activities at a community level. AEWs assume that tackling issues related to women’s and children’s nutrition to be the role of HEWs.

> *Emm… there is often a seasonal job. For example, in the health sector, they will have activities such as the* vaccination *campaign, as well as in the agriculture sector, such as spring, harvest, watershed and irrigation. We do not keep our appointment because we give priority to our main work. Especially in health, there are increased immunization programs, increased follow up programs. AEW, SNNPR*.

Integrating agriculture and health services is challenged by shortage of workforce. Food and nutrition-related interventions are not a priority for the agriculture sector. The leadership does not prioritize nutrition, and they always focus on campaigns that aim to achieve a sector-specific activity like vaccination in the health sector and water shedding and plantation in the agriculture sector. Extension workers assume that nutrition activities require additional experts assigned to the job. The service was also compromised by the absence of continuous monitoring and evaluation mechanisms.

> *The main reason was being overburdened or having overlapping duties on our side. The agricultural extension* workers *push these SURE program activities on us, by mentioning that it is not their duty. We also mentioned that these SURE program duties are their responsibility. Both sectors push the duties to each other because of work overload. Therefore, we emphasize only a single duty expected from our major career and letting others perform the SURE duties. We met and discussed the issues only when stakeholders from the zone came to our kebele for supervision, otherwise, it was overlooked by both of us. No one continuously monitors the progress as result, it will be forgotten. If this is not the case, we would perform it successfully in collaboration with AEW*. *HEW, Oromia*.

### Absence of adequate training, supervision, and technical support

Supportive supervision and technical support during the implementation of new program is a key factor for the success of a project. The SURE program was designed in a way that supervisors contribute continuously during the whole implementation period. The support provided during the program implementation was limited and many extension workers perceived the support and supervision provided were not adequate.

> *Let alone frequent training, if at least the concerned bodies at zone level visit our activities and provide the necessary feedback, it will motivate us for further improvement. It is pretty good if those in the zone were actively involved. As said earlier there is no problem at kebele level but at zone level, it seems an overlooked opportunity. HEW, Oromia*.

> *When the support is provided in an interrupted way, the community perceives the activities/supports are useless. If the support comes in an organized manner and reaches the kebele (for the farmers) hierarchically, the farmers did not oppose it. They will curiously take the supports and adapt to* their *home/household. But, if the support seems loose from its origin, the community considers it useless. AEW,Oromia*.

### Lesson learned and sustainability of health and agriculture intersectoral intervention

The SURE intervention packages depended on multi sectoral coordination at the local and district levels (Figure 1). However most agricultural interventions were perceived as the distribution of poultry and other agricultural materials. Although most extension workers captured the intention of the target population, as being mothers, young children and those who are most vulnerable households.

> *SURE, program covered mainly hen distribution for targeted population segments. For example, the program* distributed *up to 6 hens per household. Especially for the lowest quantiles of fathers and mothers* [socioeconomic]*, lactating mothers, and children less than 2 years of age. SURE, also supplies seed for vegetables. Training is also given for both the professional and community. AEW, Amhara*.

> *In the beginning, we provided training. After we took 15 days of training at the start of the program, we* mainstreamed *this to the level of community. Then, in all kebeles, chicken (for egg production) was purchased and provided for 20 individuals per kebele. Different productive agricultural materials like water pumps and others were given to support them in the production of fruits and vegetables in their backyard. AEW, Oromia*.

Health and agriculture extension workers jointly visited households with under two years of age children and provided age-appropriate IYCF counselling and advice on nutrition-sensitive agriculture to improve dietary diversity. Job aids have been developed with food calendars and recipe books for each region. Extension workers describe their experience of joint visit as follows:

> *we are working together on SURE program. The kebele has a man’s and women’s development team* [community-level volunteers who provide support to health and agriculture extension workers in mobilizing communities to deliver service]. *We and the health extension staff are going from house to house and teaching them how to feed their families, especially pregnant mothers, and children under two years of age, how to grow* vegetables *and animals on their limited land and use them at home without taking them to market. When we teach together, we or the agriculture extension workers through the men’s development team, teach them how to produce, what to sow, how to care for and feed their children together, how to feed nutritious food and how to feed at least six different types of food. The health extension worker shows them cooking demonstration. For example, they teach them what to mix, how to prepare and how often to feed. AEW, SNNPR*.

> *We gather women twice a month in their group to provide training. We have materials and different food items on our* demonstration *site, we show them how to prepare and feed soft foods like porridge for children. We advise women to prepare food available in the home based on their capacity. For instance, they sell eggs and buy cabbage, we advise them to avoid this habit and spend time feeding the child. HEW, Oromia*.

Mothers witnessed the service received by extension workers in their community. Although, asking mothers to come with food ingredients could be one factor that limits mothers from attending cooking demonstration sessions.

> *From HEW I have heard about the preparation of porridge from food items available at home like cereals (bean, chickpea and pea), seeds (teff, wheat, oats, millet an barley) vegetables and animal products (meat, egg, milk etc). I have also learnt about feeding the children only breast milk until the age of six months. …. From the AEW, I have learnt about chicken for baby and garden for baby. They taught us, as we must have vegetables in our garden and animal products at home. If these are not available at home, we are expected to buy from the market and prepare porridge from these variety of food items to feed children. Mother, Oromia*.

Farm demonstration gardens were established at the farmer’s training centers to facilitate coordination between the HEW managing the health posts and the AEWs working on the farms. Farm demonstration gardens were developed and managed by AEWs. Appropriate varieties of vegetables were selected according to agroecology. Harvests from the farmer’s training centers were used for cooking demonstrations. Cooking demonstrations were managed by HEWs at health posts near the farmer’s training centers. Women also donate ingredients from their homes to contribute to the exercise.

> *We give* agricultural *demonstrations on every occasion. Whenever we get the best performance, we give training and demonstrations on farmer’s land or on farmer’s training centers to act as the best performed farmers. We also give demonstrations as per the farmers’ needs and questions. SURE, the program recommends demonstration every month. We give porridge demonstration for both women and men developmental army every month. Before a year this was done every month consistently. Currently we give demonstrations every 2 months or longer. AEW, Amhara*.

> *When they advised us, they said in recent days, even not the rural, the urban communities are practicing* home *gardening with the limited land they have in the backyard. Different types of vegetables (tomatoes, kale, potatoes…) are on your hand; you may not need to buy them from the market, plant them. Mother, Oromia*.

### Knowledge gained through the SURE program

The SURE program interventions have impact on the knowledge of community to improve child caring practice, agricultural and behavioral practices in different parameters like severe undernutrition workload, women empowerment, and father’s support. The extension workers have observed the changes in the community, mainly fathers’ role in supporting mothers in childcaring and the importance given to feeding nutritious diet to children instead of selling the food produced. The extension workers felt that severe cases of undernourished children visiting heath centers have decreased.

> *…after the* implementation *of the SURE program, we noticed a significant decline in the prevalence of malnutrition. The number of children brought to our health post with malnutrition has significantly declined. Before the implementation of this program, we found a lot of cases and referred them. If in case we found a child with malnutrition, it is the moderate type of malnutrition. The severe form of malnutrition is almost rare after the SURE program*.

> *Concerning women empowerment, one of the most important changes SURE bring is, engaging husbands in food demonstration, letting husbands start to think as they are responsible for childcare, for their women and they also believe that women have an equal right to decide on their asset. The surprising thing was we as our area discuss many issues related to women, including their right, ability, responsibility and how they can be independent economically and manage their home. For instance, if she generated income by breeding chicken, she could obtain money on her own and incorporated it into the household after counselling and significant improvement have been observed. HEW, Oromia*.

> *In the past, men* thought *that caring for children was a women’s job. But now it has completely changed; men help their wives by caring for their children. AEW, SNNPR*.

### Importance of monitoring and regular supervision

Awareness creation activities should be organized frequently for proper implementation of nutrition interventions, and ideally by another person responsible for coordinating nutrition activities at community level. Additionally, the importance of regular meetings and evaluation for integrating services at community level was highlighted.

> *The first is, giving awareness creation education. For this HEWs and AEWs are responsible. But HEWS and AEWs are busy with* other *tasks. Therefore, there should be other responsible body at kebele level for this task to reduce the work overload and to avoid task overlapping among HEWs and AEWs. This may increase the effectiveness of the SURE program*.

> *The other is having regular meetings to monitor and evaluate the progress of the program. During this meeting, all responsible bodies should be present. The program should have a budget for both monitoring and evaluation as* well *as for supplies. The porridge and the like demonstration need raw materials. These materials should be funded by the SURE program. AEW, Amhara*.

> *At the start of the* program *there was an evaluation session every six months and sometimes every year. Now it has totally stopped. Especially after COVID 19. There is a plan, but the achievement is not checked. The reason is usually lack of budget. Therefore, there should be a budget for the monitoring and evaluation. HEW, Oromia*.

## Discussion

This study explored the experience of integrating health and agriculture sectors for improved food security and nutritional status in Ethiopia. This study reported that the major cause of undernutrition in the SURE program intervention areas are drought, conflict, and population pressure. Furthermore, the high burden of infectious diseases and WASH problems were the cause of the undernutrition. The finding is aligned with the study of Mohamed (2017) which found drought, population pressure, and conflict as the major causes of food security problems in Ethiopia, resulting in child undernutrition and household food insecurity [19]. Similarly finding on child growth in the time of drought by Hoddinott and Kinsey (2001) has shown that conflict and drought increase stunting prevalence among young children in poor households [20].

Nutrition interventions require intensive effort from multiple sectors. The expansion of governmental services from nutrition-specific to nutrition-sensitive strategies is the preferred approach, as it addresses all determinants of maternal and child nutrition and health including social and economic determinants [15]. Ethiopia has a national structure for multi sectoral nutrition coordination anchored at the Ministry of Health and a working model at district level also exists [14]. Nevertheless, according to this finding nutrition has been perceived as the work of the Ministry of Health and has been framed as a treatment/preventive problem, which mainly involves health sector interventions. One of the reasons to have a multisectoral nutrition policy is to lift the responsibility of improving nutrition into a national concern. SURE is one of the government-led programs that try to bring the health and agriculture sectors to work jointly for improved dietary diversity and improved nutritional status [17, 21]. Integrating nutrition into the national development agenda, ensuring all sectors have a mutual understanding of the importance of undernutrition prevention, and having a mechanism that helps to ensure local delivery of nutrition services, through decentralization are key components of multisectoral response for undernutrition reduction. SURE, is a common umbrella that brings HEWs, AEWs and women to promote dietary diversity and better nutrition in Ethiopia. But we found extension workers are overloaded by different sectoral activities which can limit their joint efforts and compromise the integration of nutrition-related activities. Similar to this study, frontline workers in Uganda confront various pressures from the institutional and socio-political environment, that limit their decisions about their daily activities [9]. Pelletier and collaborators found that in sub Saharan African countries, institutional integration of nutrition in different sectors was challenged by the job description of staff, the lack of sectoral alignment on multisectoral nutrition objectives and integration into the planning and reporting of sector activities [14]. A systematic review exploring the integration of nutrition-sensitive and specific interventions in fragile environments shows a lack of interest in collaboration, workload and diverting to only sectoral activities as the barriers to integrating nutrition services [22].

Delivering nutrition services to the local level tends to work better in countries that have adequate decentralized structures. In Ethiopia, the vertical structure of the Ministry of Health and Ministry of Agriculture from central to Kebele level offers an opportunity to integrate nutrition services at community level. Front-line HEWs and AEWs are a critical bridge between communities and service provision systems in the health and agriculture sectors. Empowering extension workers, supporting them and responding to the challenges they face will be an important part of ensuring the sustainability of joint activities to promote proper nutrition. Therefore, appropriate training, supportive supervision and performance monitoring should be in place to ensure the successful implementation of joint programs in the health and agriculture sectors.

### Strengths and limitations

Our study collected data from actual implementers and beneficiaries of joint intervention program which can be used to scale up their experience. However, our study focused only on the health and agriculture sectors and did not explore the experiences of other sectors included in the Ethiopian national nutrition program.

## Conclusion

Integrating existing community-level platforms are key entry points to address undernutrition and promote key agriculture and health interventions in Ethiopia. Community-based joint interventions are perceived as appropriate approaches to prevent severe cases of undernutrition and improve the role of family members in supporting mothers. Empowering extension workers through training, monitoring, and supervision, and addressing the challenges they face are part of ensuring the sustainability of integration between sectors.

### Author contributions

Conceptualization – GAM, SDH, SA

Data curation – GAM, EZT, MW

Formal analysis – GAM, SA

Investigation – GAM, MW, EZT, MW, TK, BW

Methodology – GAM, SDH, SA, TK, BW, WGB

Project administration – GAM, SDH, SA, EZT, MW

Resources – SDH, SA

Supervision – GAM, SA, EZT, MW, TK, BW

Validation – GAM, SA

Visualization – GAM, SA

Writing - original draft preparation – GAM

Writing – review and editing – GAM, SDH, SA, EZT, MW, WGB, BW, TK

## Data Availability

Data used in this analysis are available at http://data.qdr.syr.edu/

## Acknowledgment

We are grateful to the study participants. In addition, our appreciation goes to the health officials from central to community level, health, and agriculture extension workers for facilitating the implementation of the study. Finally, we would like to thank our data collectors and supervisors for their support.

## Supporting information

**S1 File. Interview guide**

**S2 File. COREQ (Consolidated criteria for REporting Qualitative research) Checklist**

